# MOLECULAR EPIDEMIOLOGY TO UNDERSTAND THE SARS-CoV-2 EMERGENCE IN THE BRAZILIAN AMAZON REGION

**DOI:** 10.1101/2020.09.04.20184523

**Authors:** Mirleide Cordeiro dos Santos, Edivaldo Costa Sousa, Jessylene de Almeida Ferreira, Sandro Patroca da Silva, Michel Platini Caldas de Souza, Jedson Ferreira Cardoso, Amanda Mendes Silva, Luana Soares Barbagelata, Wanderley Dias das Chagas, James Lima Ferreira, Edna Maria Acunã de Souza, Patrícia Louise Araújo Vilaça, Jainara Cristina dos Santos Alves, Michelle Carvalho de Abreu, Patrícia dos Santos Lobo, Fabíolla da Silva dos Santos, Alessandra Alves Polaro Lima, Camila de Marco Bragagnolo, Luana da Silva Soares, Patricía Sousa Moraes de Almeida, Darleise de Souza Oliveira, Carolina Koury Nassar Amorim, Iran Barros Costa, Dielle Monteiro Teixeira, Edvaldo Tavares da Penha, Delana Andreza Melo Bezerra, Jones Anderson Monteiro Siqueira, Fernando Neto Tavares, Felipe Bonfim Freitas, Janete Taynã Nascimento Rodrigues, Janaína Mazaro, Andreia Santos Costa, Márcia Socorro Pereira Cavalcante, Marineide Souza da Silva, Guilherme Alfredo Novelino Araújo, Ilvanete Almeida da Silva, Gleissy Adriane Lima Borges, Lídio Gonçalves de Lima, Hivylla Lorrana dos Santos Ferreira, Miriam Teresinha Furlam Prando Livorati, André Luiz de Abreu, Arnaldo Correia de Medeiros, Hugo Reis Resque, Rita Catarina Medeiros Sousa, Giselle Maria Rachid Viana

**Affiliations:** Laboratory of Respiratory Viruses, Evandro Chagas Institute (IEC), National Influenza Center (NIC) for the World Health Organization (WHO), Health Surveillance Office and Brazilian Health Ministry, Ananindeua, Pará, Brazil; Virology section, Evandro Chagas Institute (IEC), Health Surveillance Office and Brazilian Health Ministry, Ananindeua, Pará, Brazil; Laboratório Central de Saúde Pública do Acre – (Central Laboratory of Public Health of Acre). (LACEN-AC) Rio Branco, AC, Brazil; Laboratório Central de Saúde Pública do Amapá – (Central Laboratory of Public Health of Amapá). (LACEN-AP). Macapá, AP, Brazil; Laboratório Central de Saúde Pública do Amazonas – (Central Laboratory of Public Health of Amazonas). (LACEN-AM). Manaus, AM, Brazil; Laboratório Central de Saúde Pública do Pará – (Central Laboratory of Public Health of Pará). (LACEN-PA). Belém, PA, Brazil; Laboratório Central de Saúde Pública do Maranhão – (Central Laboratory of Public Health of Maranhão). (LACEN-MA). São Luís, MA, Brazil; Virology Postgraduate Program, Evandro Chagas Institute, Para, Brazil; Secretaria de Vigilância em Saúde, Ministério da Saúde – (Health Surveillance Office, Health Ministry). Brasília, DF, Brazil

**Keywords:** SARS-CoV-2, COVID-19, Amazon Region, Brazil

## Abstract

The COVID-19 pandemic in Brazil has demonstrated an important public health impact, as has been observed in the world. In Brazil, the Amazon Region contributed with a large number of cases of COVID-19, especially in the beginning of the circulation of SARS-CoV-2 in the country. Thus, we describe the epidemiological profile of COVID-19 and the genetic diversity of SARS-CoV-2 strains circulating in the Amazon Region. We observe an extensive spread of virus in this Brazilian site. The data on sex, age and symptoms presented by the investigated individuals were similar to what has been observed worldwide. The genomic analysis of the viruses revealed important amino acid changes, including the D614G and the I33T in Spike and ORF6 proteins, respectively. The latter found in strains originating in Brazil. The phylogenetic analyzes demonstrated the circulation of the lineages B.1 and B.1.1, whose circulation in Brazil has already been previous reported. Our data reveals molecular epidemiology of SARS-CoV-2 in the Amazon Region. These findings also reinforce the importance of continuous genomic surveillance this virus with the aim of providing accurate and updated data to understand and map the transmission network of this agent in order to subsidize operational decisions in public health.

## INTRODUCTION

Coronavirus disease 2019 (COVID-19) is an infectious disease caused by a newly discovered *Betacoronavirus*, now recognized as severe acute respiratory syndrome coronavirus 2 (SARS-CoV-2)^1,2^, and is responsible for one of the most significant pandemics in this century, causing millions of cases and high rates of hospitalizations and deaths^3,4^.

In South America, Brazil holds the first place of infected individuals, with 3,761,391 diagnosed cases and, out of these, approximately 118,649 Brazilians have lost their lives due to COVID-19^3^. To this end, the Amazon region has effectively contributed to the number of 760,394 infected patients and 19.358 deaths (Update August 28^th^2020)^5^ with some of the states in this region presenting the worst scenario of cases at the beginning of the pandemic in Brazil with high rates of occupancy in intensive care units (ICU) and deaths. Most persons with COVID-19 experience mild to moderate respiratory symptoms and recover^6^. On the other hand, individuals with underlying medical conditions, such as cardiovascular disease, diabetes, chronic respiratory diseases and cancer are more likely to be severely and possibly in need of intensive care ^6,7^.

In addition to epidemiological information, the SARS-CoV-2 genomic data, as well as evolution datasets to quantify the impact of non-pharmaceutical interventions (NPIs) in virus spatiotemporal spread, are under much investigation, and until then it has been shown that this virus has diversified into several phylogenetic strains^8^, marked by different punctual mutations that reflect ongoing transmission currents^9^.

In view of the above, investigations aimed at evaluating the circulation dynamics, genetics and evolutionary characteristics of SARS-CoV-2 are of substantial importance for the global surveillance of this virus and, consequently, will provide a better understanding of the virus, the disease it causes and its circulation, providing relevant information for the development of new therapeutic and control strategies and prevention of infections by the new coronavirus. To this end, we combine genetics and epidemiological data to investigate the genetic diversity, evolution and epidemiology of SARS-CoV-2 in the Amazon region.

## RESULTS

### Epidemiological data

In the Brazilian Amazon region, 8,203 samples were analyzed and 4,400 of which (53.64%) were positive for SARS-CoV-2. The frequency of detection within the region is shown in figure 1. As for circulation, the highest rate was in epidemiological weeks (EW) 17, 18, 19 and 21, with the highest peak in EW 18 (figure 2).

**Figure 1.**
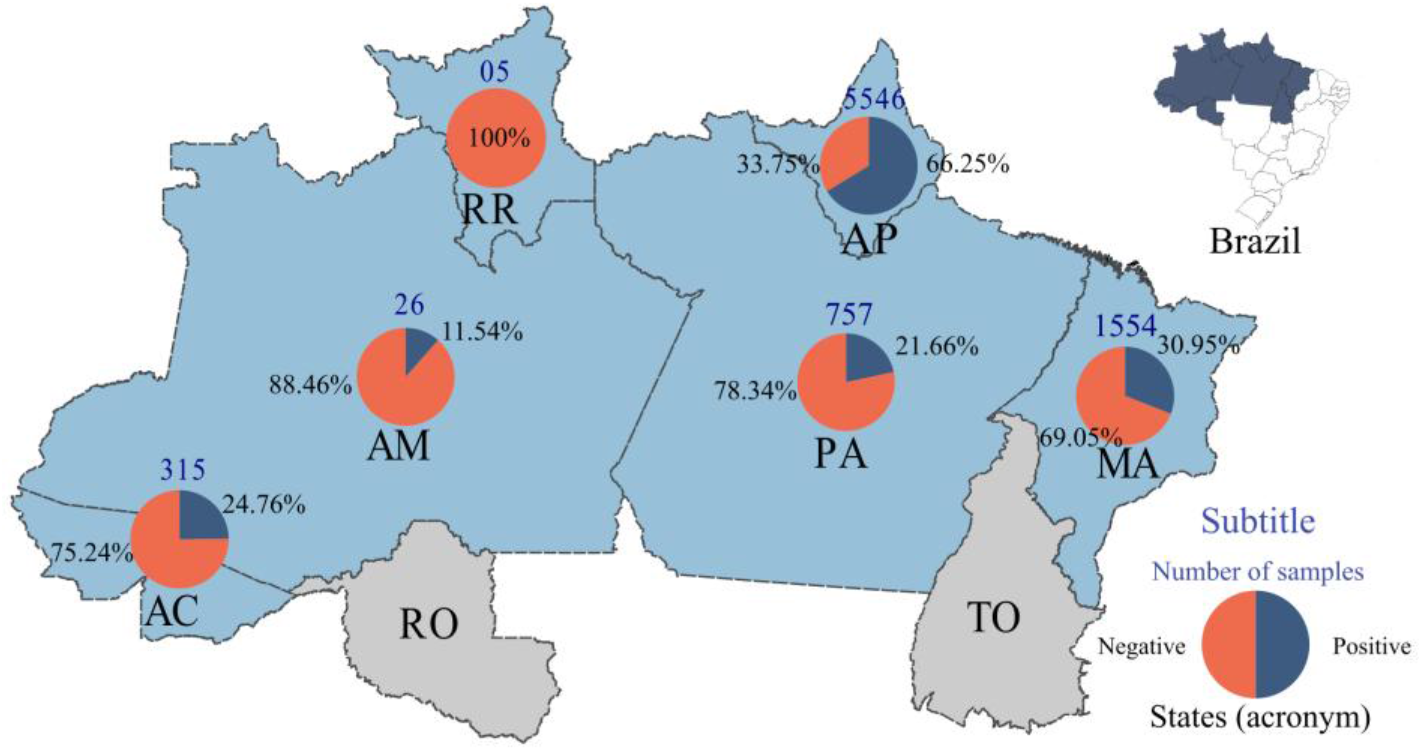
Distribution of SARS-CoV-2 cases in the Amazon region. Total number of samples studied = 8,203.

**Figure 2.**
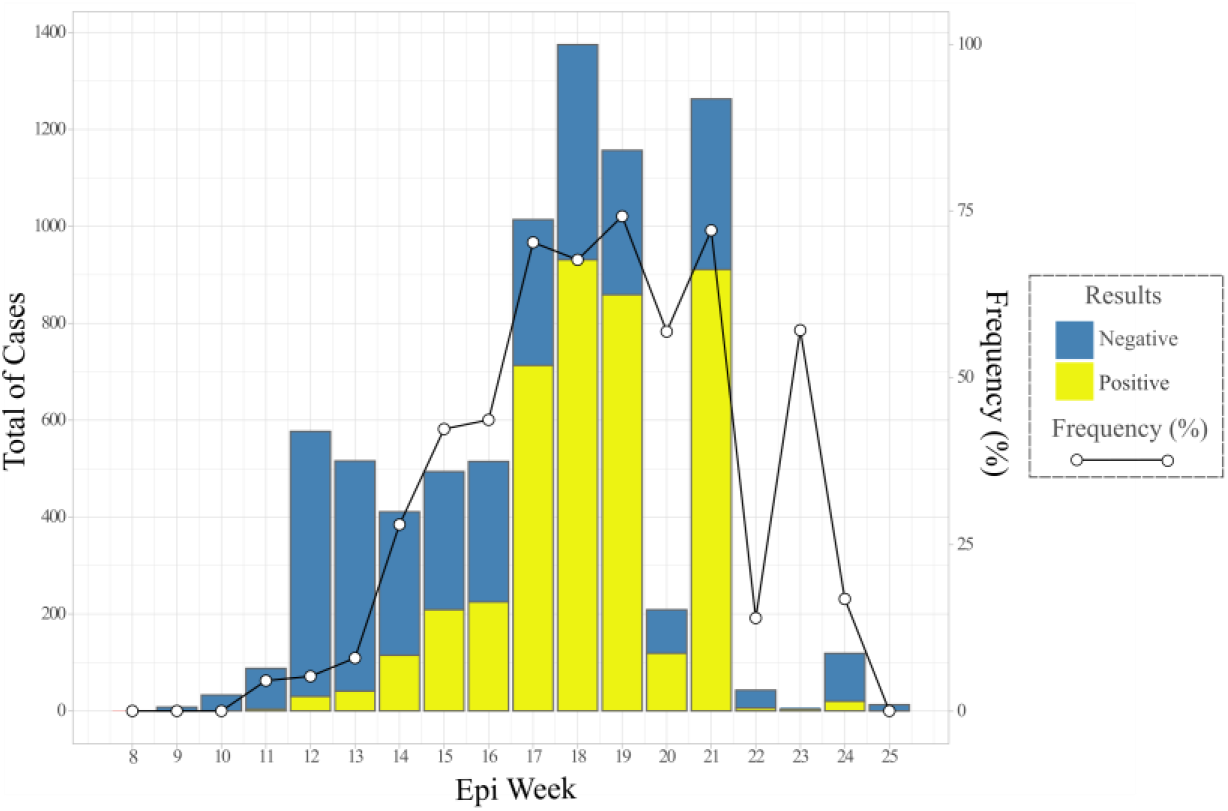
Distribution of SARS-CoV-2 cases according to the epidemiological week.

Amongst the 4,400 SARS-CoV-2 positive cases, 214 did not contain age information. In this regard, the distribution of positive samples by age group has demonstrated that the highest frequency of positivity has occurred in the adult population amongst the over 20 age groups; the average age was 47 (figure 3). Regarding sex, 2,273 (51.57%) are female and 2,116 are male (56.04%) and 11 (0.25%) did not inform it. Fever (63.11%) was the most common symptom amongst patients, followed by cough (60.70%), dyspnoea (39.52%) and sore throat (39.45%) (figure 4).

**Figure 3.**
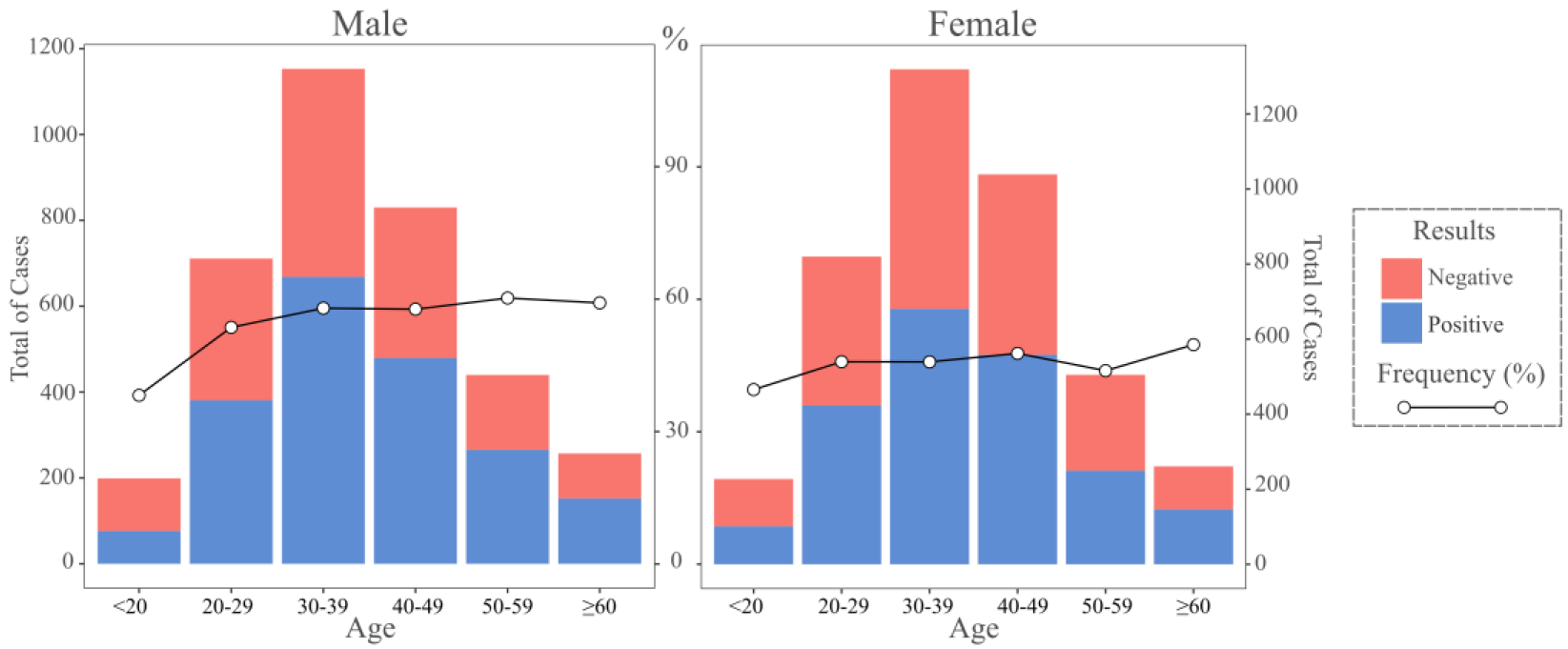
Absolute and relative frequency of positive and negative cases for SARS-CoV-2, by sex and age group.

**Figure 4.**
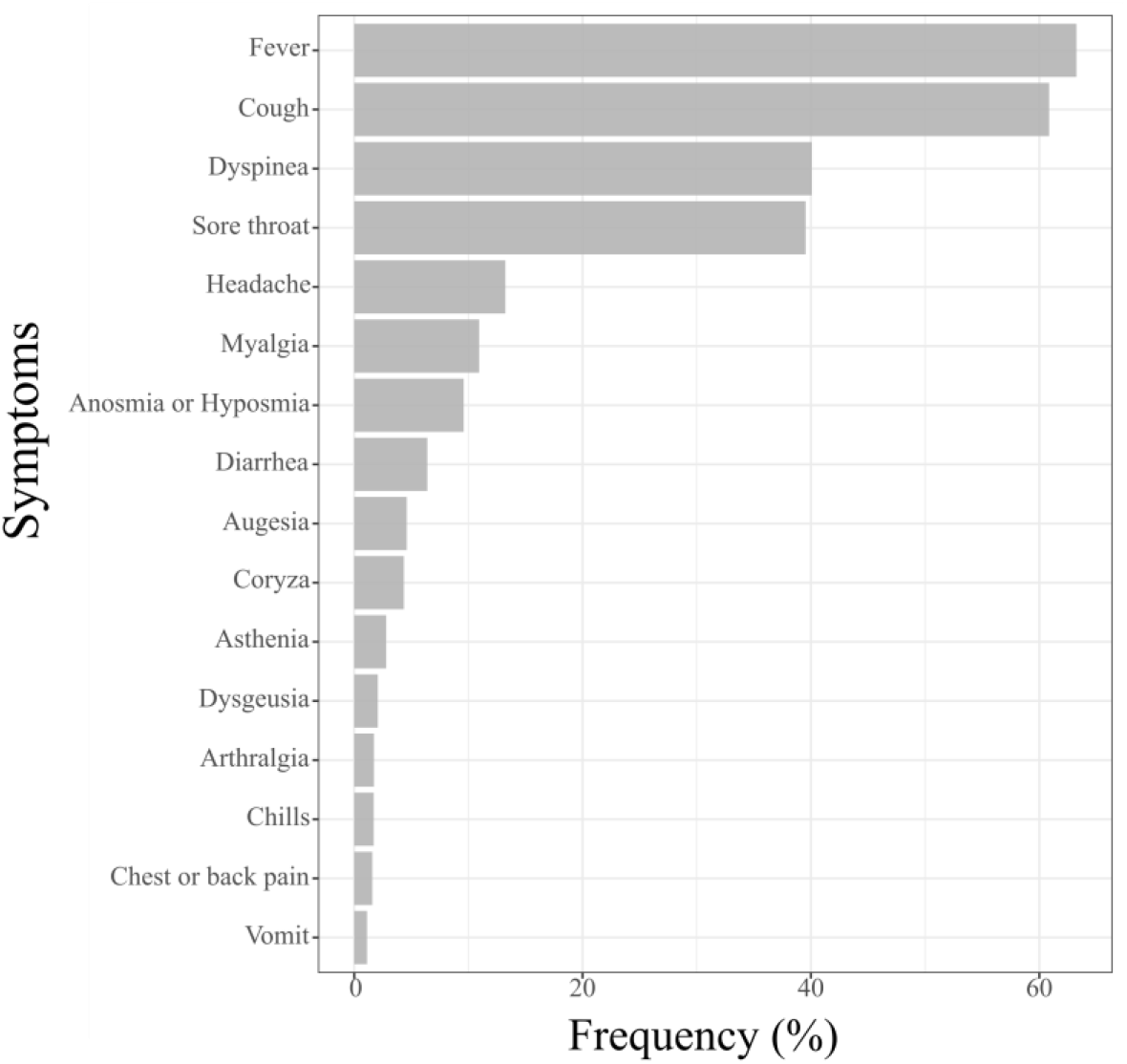
Description of symptoms and signs among positive cases for COVID-19.

Amidst the confirmed cases of COVID-19 by molecular assay, it was observed that the detection range varied between the 1st and the 42nd day after the onset of symptoms, being the fifth day (62.22%) the best collection day for detection by RT-qPCR after the onset of symptoms (figure 5).

**Figure 5.**
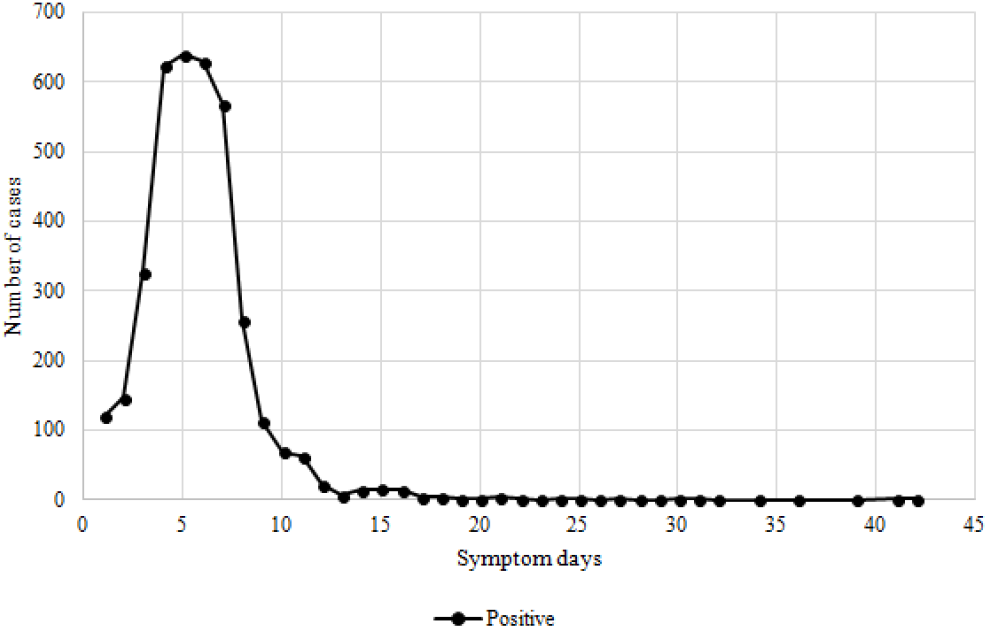
Positivity for SARS-CoV-2 regarding disease duration.

### NGS sequencing

Thirty-three (33) samples were successfully sequenced from the states of Acre (1), Amapá (11), Maranhão (8), Pará (11), Paraíba (1) and Rio Grande do Norte (1). These samples were analyzed and showed an average of 22,594,487 reads per sequenced sample, ranging from 1,476,498 to 50,601,712 reads (supplementary material 1).

### Pre-processing data

After the trimming process of regions with low quality, removal of the adapters and reads smaller than 40 bp, these samples had presented an average of 18,207,316 reads per sample, extending from 971,498 to 42,083,456 reads and then were used for assembly by De Novo and Reference Mapping (figure 6 and supplementary material 1).

**Figure 6.**
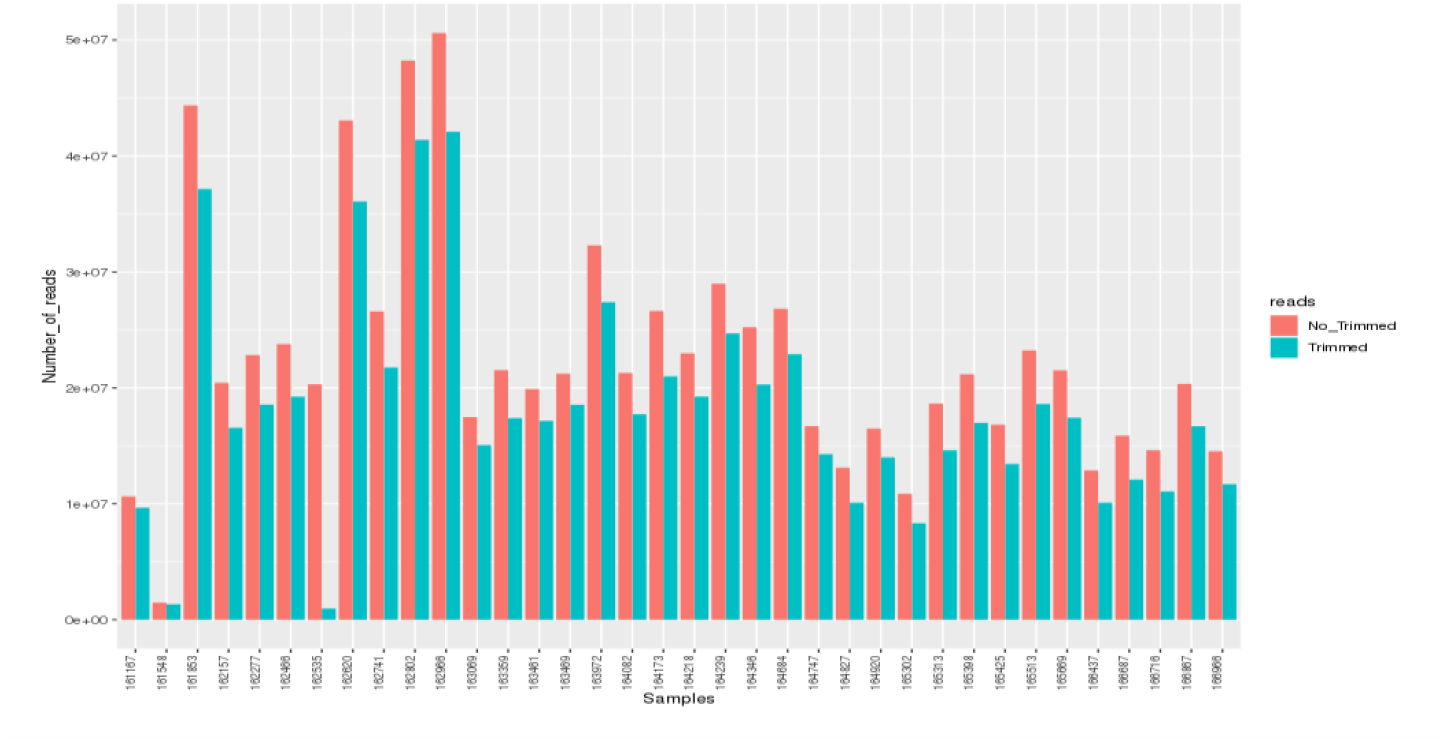
Number of reads before and after pre-processing. The reads with length less than 40 bp and quality less than Phread 20 were removed.

### Genome Assembly (De Novo and Reference Mapping)

For the genomic assembly process, the assembly was first done via the De Novo method, which has generated an average of 364,488 contigs per sample, extending from 3,964 to 1,816,455 contigs. As for the minimum size of the generated contigs is 200 bp, all the sequences under this length were discarded, for there is no variation in this item. The average of the maximum length of the contigs was 44,130 bp, extending from 4,574 bp to 312,642 bp. The generated N50 lengths were on average 409 bp, ranging from 344 bp to 618 bp (supplementary material 1).

For reference mapping assemblies, the average of reads mapped was 479,937 reads, extending from 11,935 to 3,087,036 reads per sample, leading to an average coverage of 3,007x, extending from 115x to 36,460x (figure 7 and supplementary material 1). All genomes had 38% GC content. There is no variation in this regard. All SARS-CoV-2 genomes were assembled almost entirely during assembly by De Novo, with only the ends needing editing, assembled via reference mapping and later sequence edition.

**Figure 7.**
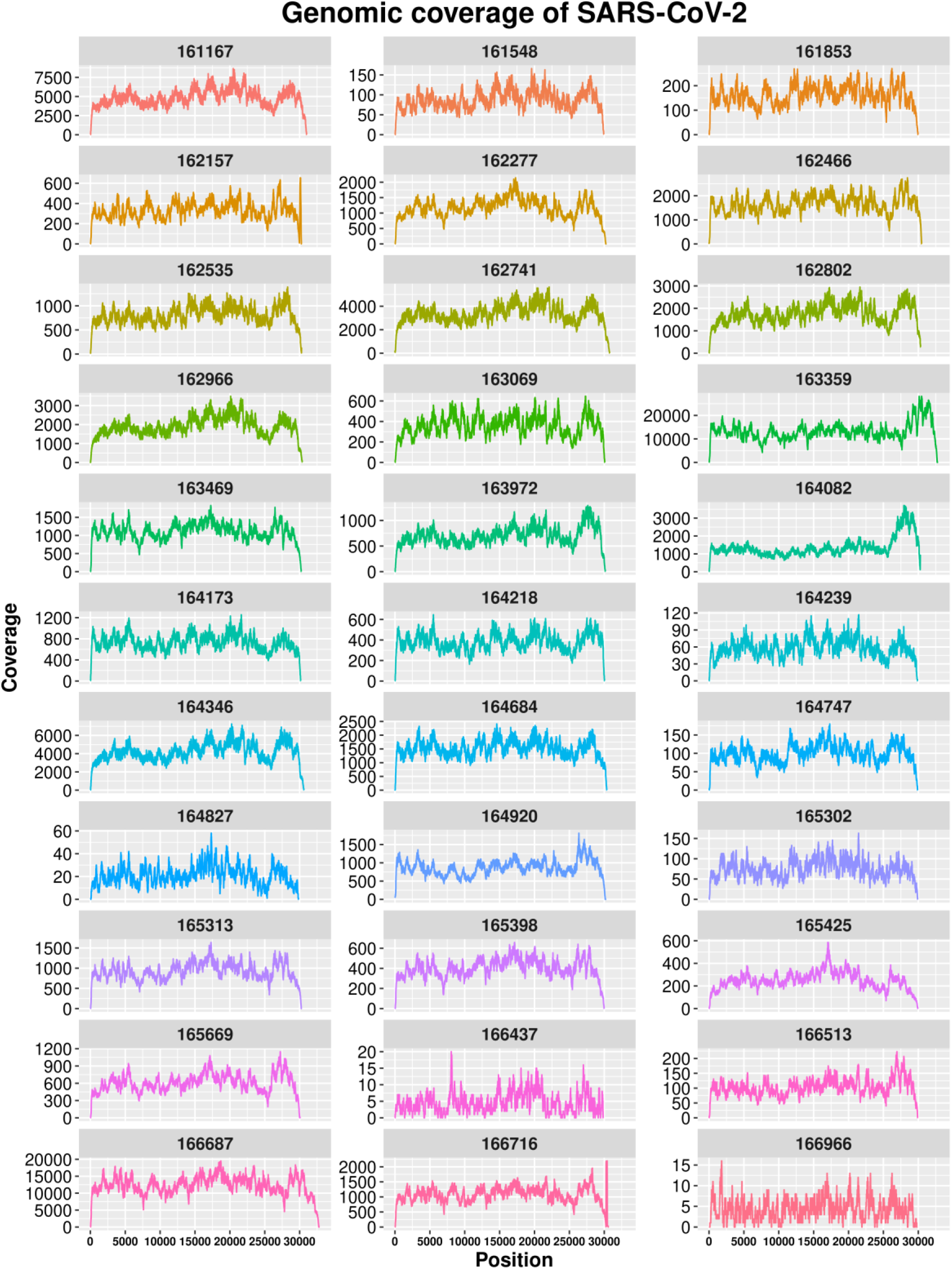
Coverage plot by sample sequenced in the present study.

### Phylogenetic Analysis and Mutation analysis

The genomes obtained were aligned with the reference strains deposited in GISAID, showing an identity of 99.98% (supplementary material 2). The analysis of the SARS-CoV-2 genome found revealed 62 nucleotide changes in 12 genes leading to 32 amino acid changes in 7 proteins (supplementary material 3).

The phylogenetic analysis reveals that isolates from present study clustering in three major clades in B.1 (one clade) and B.1.1 (two clades), with moderate statistical values of 70-89%. These clades are characterized by the presence of mutations S:D614G (B.1 and B.1.1) and substitutions in protein N (N:R203K and N:G204R) that classifies the lineage B.1.1 (Figure 08).

**Figure 8.**
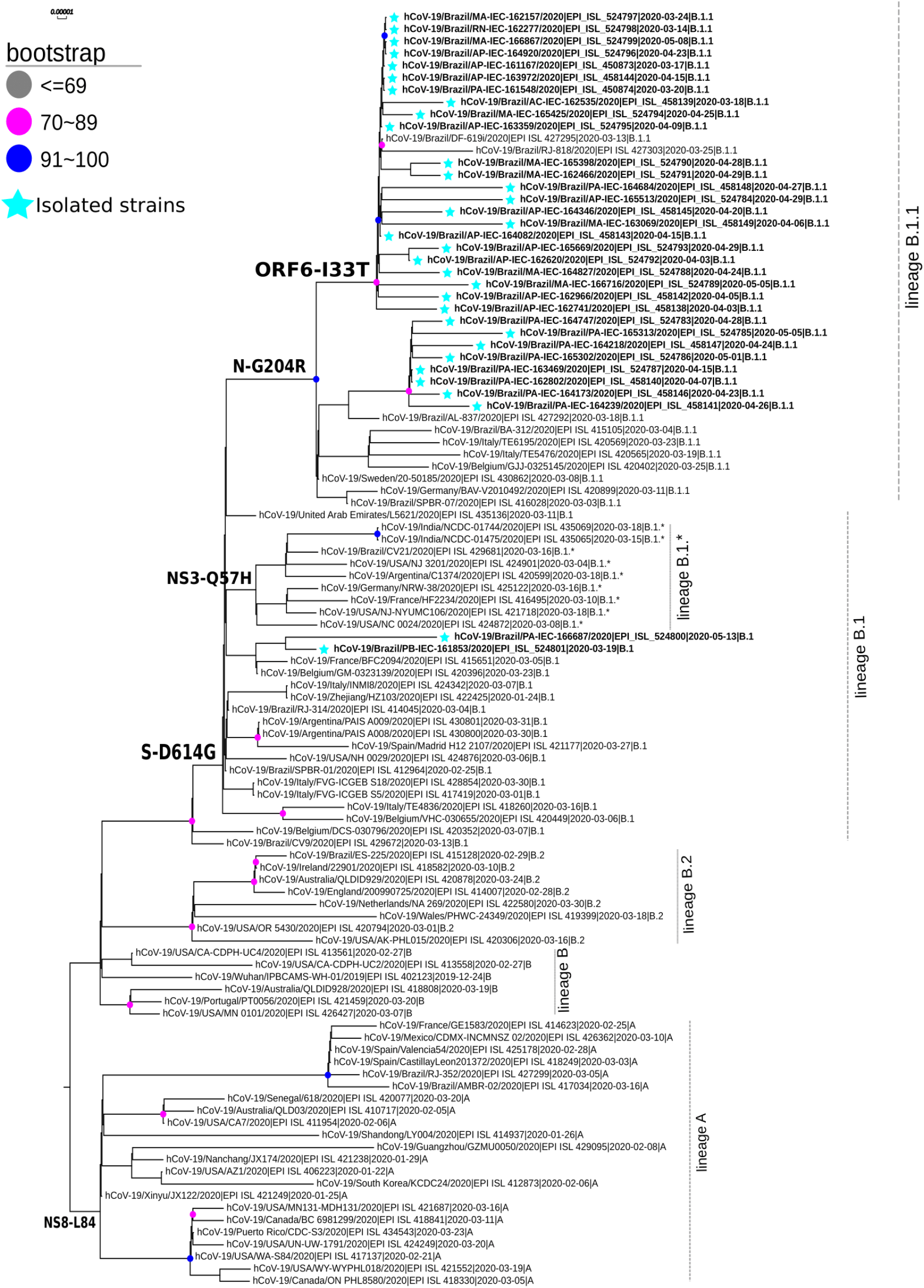
SARS-CoV-2 complete genome phylogenetic tree (ML) with 1000 bootstraps, using GTR evolution model for nucleotide substitutions.

## DISCUSSION

In Brazil, the first case of COVID-19 was detected in the state of São Paulo on February 26, 2020. In the Brazilian Amazon Region, the first detection has occurred in the state of Manaus on March 13, 2020 and, in that month, the Brazilian Health Ministry had already reported community transmission in the country, as well as a pandemic status by the World Health Organization (WHO). After the first detection of SARS-CoV-2 in this Brazilian site, an extensive spread of this virus was observed in the region, demonstrating, in the present study, a detection frequency of 53.64% showing a critical adaptation and circulation of SARS-CoV-2 in this tropical region. The most active circulation of SARS-CoV-2 has occurred mainly in April and May, in EW 17, 18, 19 and 21, which coincides with the Amazonian winter season.

The detection of SARS-CoV-2 has occurred in all age groups. However, the highest frequency has occurred in adults and the elderly, as described in the world population^10-13^. The low frequency of SARS-CoV-2 in children and teenagers under 20, verified in the present study, has been associated with reduced susceptibility and less likelihood to infection or a combination of both, compared to adults^14-16^.

As for sex, similar to what has been reported by literature, the frequency amongst men was slightly higher than in women^17,18^. Estrogen, the main female sex hormone, plays a possible protective role in COVID-19, activating the immune response, and directly suppressing SARS-CoV-2 replication^19,20^. Indeed, estrogen inhibits the activity or expression of different components of the renin-angiotensin system. Particularly, estrogen can upregulate ACE2 expression^21,22^. Regarding the clinical analysis presented by the investigated patients, the most described symptoms were fever, cough, sore throat and dyspnoea, respectively, associated or not, commonly reported amongst respiratory infections, as well as in COVID-19^10,11,23,24^. However, unlike other respiratory infections, in COVID-19 anosmia and dysgeusia have been frequently reported^25-29^, as described amongst patients in the study region. After the onset of symptoms, the period of greatest detection of SARS-CoV-2 by RT-qPCR has occurred on the fifth day, similarly to what has been described in other studies^30^.

The genomic analysis of the viruses found revealed that 61 nucleotide mutations were found in the entire genome when compared to the reference genome, and, out of these, 16 led to amino acid changes, with emphasis on the substitutions in S and N proteins that have a structural role and ORF6, a non-structural protein not yet characterized ^31,32^.

Amongst the alterations in the Spike protein that plays a role in binding to the human ACE2 receptor and is also the main antigenic target, it was found the D614G substitution that is described as a factor that antigenically favors the virus, giving it a higher capacity to infection^33^ and has been used as a genetic marker for strains of the B-lineage (Pangolin Classification) which has become the largest circulating group worldwide^34^. Also, it was verified the V1176F mutation described in the literature^35^ and used as a genetic marker for samples circulating in Brazil (https://www.gisaid.org), but no antigenic advantage has yet been attributed.

Regarding the N protein plays a role in folding viral genetic material and has been used as a marker for samples from Europe, it was verified R203K, G204R and I292T amino acidic substitutions. However, their molecular roles are still unclear^36,37^. The change I33T in ORF6, a non-structural protein, has been observed in samples originating in Brazil and that circulate in South America^38^.

The phylogenetic analysis revealed that the samples of this study have formed three distinct groups that cluster with the phylogenetic lineages B.1 and B.1.1 that have samples already sequenced from Brazil^39^. Within clade B.1, only two samples from Pará were clustering with samples from Europe. In clade B.1.1, it was possible to observe the formation of two distinct groups divided by the I33T ORF6 and V1176F S protein substitutions. These two mutations have been observed to divide the two main strains of SARS-CoV-2 circulating in Brazil^34^. Since its worldwide circulation on December 2019^40^, the SARS-CoV-2 genome has changed wherever it arrives^41^, which may mean a likely adaptation to the population^42^.

In this study, we did not yet had the chance to analyze how SARSCoV-2 became established across the Amazon region and to associate the finding lineages with the population movements, that is, to relate to the proportion of within and between state measured virus movements. Another relevant issue is that the B.1 and B.1.1 lineages from the Amazon region were quite similar, making it difficult to trace with precision the origin of these strains in the study site.

In conclusion, this study reveals that the highest SARS-CoV-2 circulation has reached its peak in epidemiological week 18. The distribution of positive samples by age group has demonstrated that the median age was 47, with men being the main affected gender and there was a spectrum of symptoms composed of fever, cough, dyspnoea and sore throat. Furthermore, this investigation supports the evidence for the existence of two main lineages (B.1 and B.1.1) associated with genomic epidemiology of SARS-CoV-2 in the Amazon region. Thus, genomic surveillance must be continuously adopted to be able to offer accurate and quality data to understand where this virus emerged from, and map the transmission network to improve operational decisions in public health.

## METHODS

### Samples and ethical aspects

The Laboratory of Respiratory Viruses of the Evandro Chagas Institute (LVR-IEC), located in the Amazon region, works with the World Health Organization (WHO) as a National Influenza Center (NIC) for the surveillance of influenza and other respiratory viruses, amongst them, the SARS-CoV-2. Thus, this laboratory has received 8,203 clinical specimens from patients of both sexes and in different age groups (zero to 111 years old) between February 27^th^, 2020 to July 1^st^, 2020 for the diagnosis of SARS-CoV-2 from the states of Acre, Amapá, Amazonas, Ceará, Maranhão, Pará, Paraíba, Pernambuco, Rio Grande Norte and Roraima. The clinical specimens collected and used for molecular diagnosis and viral genetic analysis were nasopharyngeal swabs plus throat swabs, nasopharyngeal aspirate and sputum. This study was approved by Evandro Chagas Institute Ethical Committee (34931820.0.0000.0019).

### Extraction and Detection by RT-qPCR of viral nucleic acid

The viral RNA was extracted manually using the QIAamp® Viral RNA Mini Kit (QIAGEN, Hilden, Germany) following the manufacturer’s guidelines. The detection of the viral genome by RT-qPCR was performed with the Molecular Kit SARS-CoV-2 (E/RP) Biomanguinhos (Biomanguinhos, Rio de Janeiro, Brazil), according to the protocol described by Corman et al (2020)^45^.

The amplification reaction was conducted sequentially in the following steps: reverse transcription at 50°C for 15 minutes, followed by transcriptase inactivation and activation of Taq DNA polymerase at 95 ° C for 2 minutes, polymerase chain reaction at 95°C for 15 seconds in 45 cycles, extension and annealing at 55 ° C for 30 seconds. At the end of the amplification, all clinical samples should have reaction sigmoid curves for the targets that cross the limit line *(cycle threshold* – Ct) equal to or before 40 cycles. Positive and negative controls were included in each reaction.

### Epidemiological analysis

Graphs of epidemiological data (age, sex, state, signs and symptoms) and circulation were performed with support by the LVR-IEC database and the Microsoft Office Excel program. The data were inspected, visualized and plotted using the R programming language script^43^ together with the libraries ggplot2^44^, geobr^45^, pipeR^46^, readr^47^, lubridate, fmsb^48^, plyr^49^, scales^50^, viridis^51^ and hrbrthemes^52^. By international convention, epidemiological weeks were counted from Sunday to Saturday, considering the sample collection date.

### Sample selection for sequencing

The selection of strains for sequencing the viral genome was conducted so that there was geographical and temporal representativeness. In this aspect, the date of collection and the respective epidemiological week of the sample of each state of origin were considered to reach the minimum representation of each federated unit per epidemiological week. In addition, in order to obtain the highest amount of viral RNA and, thus, a greater chance of success in sequencing, samples that showed Ct ≤ 20 in the RT-qPCR for SARS-CoV-2 were selected.

### Library construction and sequencing

The total RNA was converted to complementary DNA (cDNA) using the cDNA Synthesis System Kit and 400 μM of random primers, following the manufacturer’s procedure. The reaction solution was purified with the Agencourt AMPure XP Reagent. The cDNA library was prepared and sequenced using the methodology described in the Nextera XT DNA Library Preparation Kit on a NextSeq (Illumina, Inc) platform by paired-end methodology with 300 cycles (2×150 reads), in the Evandro Chagas Institute, Brazil Ministry of Health.

### Data pre-processing

The data were evaluated for their quality regions. The adapters sequences reads with a quality lower than Phred 20 and reads with less than 40 bp size, were removed using Trimmommatic^53^. The processed reads were visualized with FastQC^54^. For Trimmomatic, we have used the following parameters: LEADING: 3 TRAILING:3 MINLEN:40

### Genome Assembly *(De Novo* and *Reference Mapping)*

For this step, the reads validated based on quality trimming were used to assembly the SARS-CoV-2 genomes. The De Novo assembly was performed using the Megahit v.1.1.4-2^55^ and for Reference Mapping we have used the software Bowtie2^56^ and Geneious Prime, where the respective coverage, gaps and final size of the genome were analyzed. For genome assembly, all programs were performed with default parameters.

### Taxonomic annotation and submission to GISAID

The generated de novo contigs were compared using the Blastx tool^57^ implemented in Diamond v.0.9.3 3 ^58^, against the RefSeq database (NCBI’s Protein Reference Sequences Database), which is a database of cured protein sequences and which provides a high level of annotation, such as the description of the function of a protein, its domain structure, post-translational modifications, where a statistical value (e-value) of 0.0001 was considered.

The viral genome annotation was performed automatically using the Geneious Prime software *(Biomatters, Ltd., New Zealand*, 2019) and cured manually by comparing the starts and stop codons, as well as the sizes of the genes. These genome sequences were subsequently submitted to the GISAID database (https://www.gisaid.org/) under accession numbers EPI_ISL_450873-450874, EPI_ISL_458138-EPI_ISL_458149 and EPI_ISL_524783-EPI_ISL_524801.

### Phylogenetic Analysis and Mutation analysis

The genomes sequences were aligned with other genomes from all the world using the Mafft v.7.471^59^. For phylogenetic analysis the software RaXML^60^ with 1000 bootstraps was used as statistical support, using GTR as a nucleotide substitution model. The genomes obtained were compared to the reference strain (NC_045512) by *in house* python script that compares each base of the entire genome and gives us a mutation list.

## Data Availability

All data is available on GISAID

## Acknowledgements

The authors would like to thank all the professionals who worked bravely to deal with this pandemic, especially in the Amazon. We thank the Evandro Chagas Institute, where the development of the research was carried out with great contribution from the virology team. We would also like to thank the General Coordination of Laboratories (CGLab) of the Ministry of Health (MS), States of the Brazilian Central Laboratory (LACENs), and local surveillance teams for the partnership in viral surveillance in Brazil.

## Author contributions

MCS, HRR, RCMS and GMRV coordinated the study; ECSJ, JAF, AMS, SPS, MPCS and JFC performed the sequencing and genomic analysis of SARS-CoV-2 strains; LSB, WDCJ, AMS and JAF performed the detection of SARS-CoV-2 by RT-qPCR; AMS, JLF, EMAS, CKNA and DSO received and checked the samples; PLAV, JCSA, MCA, PSL, FSS, AAPL, CMB, LSS and PSMA performed the registration of epidemiological data in a database and assisted in the release of results; IBC, DMT, ETPJ, DAMB, JAMS, FNT and FBF performed the extraction of the viral genome; MSS, GAN, IAS, GALB, LGL, HLSF collected the samples; MTFPL, ALA and ACM assisted with technical support through the viral surveillance network in Brazil.

## Additional Information

The authors declare that they have no conflicting interests.

